# Low pre-infection levels of neutralizing antibody in breakthrough infections after bivalent BA.4-5 vaccine and practical application of dried blood spots

**DOI:** 10.1101/2024.01.30.24301988

**Authors:** Hitoshi Kawasuji, Yoshitomo Morinaga, Hideki Tani, Hiroshi Yamada, Yoshihiro Yoshida, Masayoshi Ezaki, Yuki Koshiyama, Yusuke Takegoshi, Makito Kaneda, Yushi Murai, Kou Kimoto, Kentaro Nagaoka, Hideki Niimi, Yoshihiro Yamamoto

## Abstract

The level of neutralizing antibodies required to confer protection against COVID-19 breakthrough infections (BIs) is unclear, and the ability to know the immune status of individuals against the rapidly changing endemic variants is limited. We assessed longitudinal serum anti-RBD antibody levels and neutralizing activities (NTs) against Omicron BA.5 and XBB.1.5 in healthcare workers following the fourth monovalent and fifth bivalent BA.4-5 vaccines. The occurrence of BIs was also followed, and pre-infection antibody levels were compared between patients who developed BI and those who did not. In addition, we collected whole blood samples on the same day as the sera and stored them on filter papers (nos. 545, 590, and 424) for up to two months, then measured their NTs using dried blood spots (DBS) eluates, and compared them with the NTs in paired sera. Pre-infection levels of NTs were lower in patients who developed BI than those who did not, but the anti-RBD antibody levels were not different between them. The NTs below 50% using 200-fold diluted sera might be one of the indicators of high risk for COVID-19 BI. However, the NTs against XBB.1.5 at 6 months after the fifth dose of bivalent BA.4-5 vaccine were lower than this threshold in almost half of infection-naïve participants. NTs measured using DBS eluates were strongly correlated with those measured using paired sera, but the time and temperature stability varied with the type of filter paper; no. 545 filter paper was found to most suitable for NT evaluation.

## INTRODUCTION

Messenger RNA (mRNA) vaccines are highly effective in protecting the population from symptomatic and severe disease, hospitalization, and death [1, 2]. Nevertheless, vaccine efficacy (VE) has gradually waned over time [3], and has further decreased with the emergence of variants of concern, including XBB.1.5 and the other Omicron XBB variants [4]. Due to waning immunity of mRNA monovalent vaccines and to protect against emerging variants, bivalent mRNA vaccines (Original/Omicron BA.1 or BA.4/BA.5) were authorized and widely used [5], but the emergence of Omicron XBB subvariants has dramatically reduced the neutralizing antibody responses of both the SARS-CoV-2 wild-type (WT) monovalent and bivalent vaccines [6]. Therefore, monovalent XBB.1.5-spike-based vaccines were developed and authorized for use in the Fall of 2023. As of May 2023, however, only 17% of the eligible US population had received the bivalent booster dose [5, 7]. This low vaccination rate may be due to not only adverse reactions to vaccinations, but also the unclear thresholds for protection against COVID-19 breakthrough infection (BI) and the limited ability to know the immune status of individuals against the rapidly changing endemic variants in real time.

Since decreased VE has been correlated with the decrease of mRNA vaccine-induced neutralizing antibodies occurring over time [8, 9], a lower level of neutralizing antibodies was assumed to be associated with a higher risk of BI. However, the association between pre-infection humoral immunity and the risk of BI during the WT and Delta-dominant periods was not clarified in the previous epidemiologic studies [10–12]. Furthermore, during the Omicron dominant period, studies investigating whether a lower level of vaccine-induced pre-infection humoral immunity is associated with a higher risk of BI remain scarce.

To investigate humoral immunity against SARS-CoV-2, many assays are used but not all assays measure neutralizing antibodies that can bind and subsequently prevent viral entry, and correlate with protection against future infections [13]. Live virus assays are the gold standard for measuring neutralizing antibody responses, but they are not suitable as a routine test in clinical laboratories due to their complexity and the risks associated with using live viruses [14]. We previously established the high-throughput chemiluminescence reduction neutralization test (htCRNT) for the evaluation of immunity to SARS-CoV-2, using a pseudotyped virus [14, 15]. Neutralizing assays using the pseudotyped virus overcome the containment problems related to use of a live virus, and the htCRNT values were found to correlate with the results obtained in live virus assays [16].

Nevertheless, SARS-CoV-2 antibody testing currently requires serum or plasma collected by venipuncture. The use of such sampling in large-scale seroepidemiologic studies is limited by resources, costs, and the risk for SARS-CoV-2 exposure from direct patient contact [17]. In contrast, dried blood spot (DBS) sampling is simple and inexpensive; the DBS samples can be self-collected and remain stable at ambient temperature until sent by postal mail to a laboratory for processing [17]. Moreover, DBS sampling has the potential to provide a widely available solution for assessing and tracking herd immunity in various populations, including low- and middle-income countries, without geographic limitation. To date, DBS sampling is best known for its use in screening newborns for genetic health conditions, and some studies have utilized DBS to quantify SARS-CoV-2 antibody levels [17, 18]. However, only a few studies have explored the use of DBS to measure SARS-CoV-2 neutralizing antibody responses [13, 19].

We conducted a prospective longitudinal study to assess the immunogenicity and durability of the third booster dose (fifth dose) with the Omicron BA.4-5 bivalent vaccine in healthcare workers. To estimate the immune threshold for protection against COVID-19 BIs, we also compared pre-infection anti–receptor-binding domain (RBD) or neutralizing antibody levels between breakthrough cases and controls during the BA.5 and XBB sub-lineages endemic period. Moreover, by comparing neutralizing activities (NTs) in paired sera to eluates from simultaneously collected DBS cards, we have shown that DBS are a viable and time- and temperature-stable sample collection method for the evaluation of neutralizing antibody responses against variants.

## MATERIALS AND METHODS

### Study design and participants

This prospective longitudinal cohort study was initiated at Toyama University Hospital at the time of the primary vaccination in 2021 [20]. All participants were healthcare workers who had received the four monovalent vaccines followed by a fifth dose of the BA.4-BA.5 bivalent vaccine (Pfizer-BioNTech COVID-19 vaccine, bivalent [original and Omicron BA.4/BA.5]) on the same schedule, and had no recorded COVID-19 infections at 3 months after the fourth dose (3mA4D). All participants were previously included in our studies on the safety and immunogenicity of the primary vaccination (first and second doses) of BNT162b2 [20], the first booster (third) dose of BNT162b2 [21], and the second booster (fourth) dose of mRNA-1273 vaccine [16]. In the present study, the longitudinal sera were subsequently collected at 3mA4D, 2 weeks after the fifth dose (2wA5D), and 6 months after the fifth dose (6mA5D) to assess humoral responses to the fourth and fifth vaccination doses. Data on age and sex of all participants in the present study were extracted from the databases from the previous studies [16, 20, 21].

Participants with previous SARS-CoV-2 infections were excluded for the evaluation of immunogenicity. Diagnosed infection was defined as a history of COVID-19 (confirmed by positive RT-PCR or antigenic test) that was self-reported, along with the presence of positive anti-SARS-CoV-2 nucleocapsid (N) antibodies. Cases without a history of COVID-19, but which developed positivity for anti-N antibodies, were defined as undiagnosed infections. The Toyama University Hospital Infection Control Team received reports from all the infected staff and their departments and gave them instructions on their return-to-work dates; therefore the date of diagnosis, diagnostic procedure, possible route of infection (close contact), symptoms, and hospitalization were recognized and recorded for all cases.

### COVID-19 BI during the study period

During the longitudinal study period from 15 November 2022 to 30 September 2023, we identified 10 diagnosed and one undiagnosed infection between 3mA4D and 2wA5D, 2 diagnosed infections between 2wA5D and 6mA5D, and 9 diagnosed infections after 6mA5D. The date of symptoms onset for all cases was between 4 December 2022 and 6 January 2022 during the period from 3mA4D to 2wA5D, between 11 and 15 January 2023 during the period from 2wA5D to 6mA5D, and between 5 June and 22 September 2023 during the period after 6mA5D. We could not conduct whole-genome sequencing for variants analysis, but the genomic survey of the local institute of public health reported that the Omicron BA.5 variant was dominant, with only a few populations of BA.4, BF.7 or BQ.1 variants, during the periods from 3mA4D to 2wA5D and 2wA5D to 6mA5D [22]. In the period after 6mA5D, XBB sub-lineages (XBB.1.5, XBB.1.9, XBB.1.16, and EG.5) were dominant [22]. Therefore, we evaluated the NTs against BA.5 and XBB.1.5 as representative variants of XBB sub-lineages.

To estimate the cut-off value of the antibody level associated with high risk of COVID-19 BI, among participants infected between 3mA4D and 2wA5D, pre-infection (at 3mA4D) anti-RBD antibody and NTs against the infected Omicron BA.5 variant were compared between BI cases and controls without BI. Similarly, during the period after 6mA5D, pre-infection (at 6mA5D) anti-RBD antibody and neutralization activities against XBB.1.5 were compared between BI cases and controls.

### Sera collection

Serum samples were collected from the participants at 3mA4D, 2wA5D, and 6mA5D. The sera were used for serological assays within three days of storage at 4°C or frozen at −80°C until further verification. In DBS evaluation, paired sera were collected on the same day as DBS.

### Dried blood spot collection and processing

DBS specimens were collected by self-collected finger-prick blood drop or blood drop of the residual whole blood dried on the no. 545, 590, and 424 filter papers (Advantec, Tokyo), which have been extensively used in newborn screening and other applications. The DBS cards were stored (i) at room temperature (RT, 26°C) overnight; (ii) at RT for one month; or (iii) at RT for one month (1M) and then at high temperature (HT, 40°C) for an additional month (two months total [2M/HT]) until analyzed. For the elution process, a 4 mm disc was punched from the center of each DBS card and incubated with 100 μL of Dulbecco’s modified Eagle’s medium (DMEM; Nacalai Tesque, Kyoto, Japan) containing 10% heat-inactivated fetal bovine serum (FBS) for 2 h at RT. The eluent was either tested immediately or stored at −80°C for long term storage. Based on the blood absorption per area, which was similar among the no. 545, 590, and 424 filter papers, the assumption that the serum accounted for approximately 50% of the whole blood, and 100 μL of medium used for elution, the eluents were considered to be equivalent to a 20-fold dilution of sera.

### SARS-CoV-2 pseudotyped virus neutralization assay

Pseudotyped vesicular stomatitis virus (VSV) containing the SARS-CoV-2 S protein was generated as previously described [15]. The expression plasmids for the truncated S protein of the SARS-CoV-2 variants, pCAGG-pm3-SARS2-SHu-d19_D614G (Wuhan), pCAGG-pm3-SARS2-SHu-d19_B.1.1.529.5 (Omicron BA.5-derived variant), pCAGG-pm3-SARS2-SHu-d19_XBB1.5 (Omicron XBB.1.5-derived variant), and VSVs bearing envelope proteins (VSV-G) were provided by Drs. C. Ono and Y. Matsuura of the Research Institute for Microbial Diseases, Osaka University, Japan. The pseudotyped VSVs were stored at −80°C until use.

The neutralizing effects of each sample against pseudotyped viruses were examined using a htCRNT as previously described [14, 16]. Briefly, serum was diluted from 100- to 25600-fold, and elution of the DBS (equivalent to 20-fold dilution of sera) was diluted from 5- to 1280-fold (equivalent to 100- to 25600-fold diluted sera) with DMEM containing 10% heat-inactivated FBS and incubated with pseudotyped SARS-CoV-2 for 1 h. After incubation, the VeroE6/TMPRSS2 cells (JCRB1819) were treated with pseudotyped viruses and DMEM-containing serum or elution of DBS. The infectivity of the pseudotyped viruses was determined by measuring the luciferase activity after 24 h of incubation at 37°C. The infectivity of samples without pseudotyped virus was defined as 0% infection, and that of pseudotyped virus without serum and elution of DBS was defined as 100% infection (100% and 0% inhibition), respectively.

### Anti-RBD and anti-nucleocapsid antibody measurements

The concentration of anti-RBD antibodies in serum samples was measured using the Elecsys Anti-SARS-CoV-2 S immunoassay (Roche Diagnostics GmbH, Basel, Switzerland) at Toyama University Hospital. At 3mA4D, 2wA5D, and 6mA5D, most serum samples exceeded the upper limit of quantification (25000.0 U/mL); therefore, these samples were diluted 4-fold manually before measurement. Measurements of >100000.0 U/mL were considered 100000.0 U/mL for further statistical calculations. The serum concentration of anti-N antibodies was measured using an Elecsys Anti-SARS-CoV-2 immunoassay (Roche Diagnostics GmbH, Basel, Switzerland). The anti-N antibody levels were expressed as a cut-off index value; values ≥1.0 were considered positive for anti-nucleocapsid antibodies.

### Statistical analysis

Statistical analysis was performed using the Mann–Whitney test to compare non-parametric groups. Correlations between the test findings were expressed using Pearson’s correlation coefficient. Data were analyzed using GraphPad Prism version 9.5.1 (GraphPad Software, San Diego, CA). Statistical significance between different groups was defined as *P* <0.05. Data are expressed as mean with standard deviation (SD) or median with interquartile range (IQR).

### Ethics approval

This study was performed in accordance with the Declaration of Helsinki and approved by the ethical review board of the University of Toyama (approval no. R2019167). Written informed consent was obtained from all participants.

## RESULTS

### Study flow chart

For the longitudinal evaluation of participants with a known vaccine history and known immune status, all participants who received four monovalent vaccines and a fifth dose of the BA.4-BA.5 bivalent vaccine on the same schedule were eligible [16, 20, 21]. Anti-RBD antibodies and NTs against Omicron variants BA.5 and XBB.1.5 were initially measured in 133 participants at 3mA4D (**Fig. 1**). After the third booster vaccination (fifth dose), the humoral immunity of participants was longitudinally assessed at 2wA5D and 6mA5D. Participants with previous SARS-CoV-2 infections were excluded from the longitudinal study to assess the immunogenicity of the third booster dose with the Omicron BA.4-5 bivalent vaccine in healthcare workers. The demographic characteristics of the participants are shown in **Table 1**.

**Figure 1.**
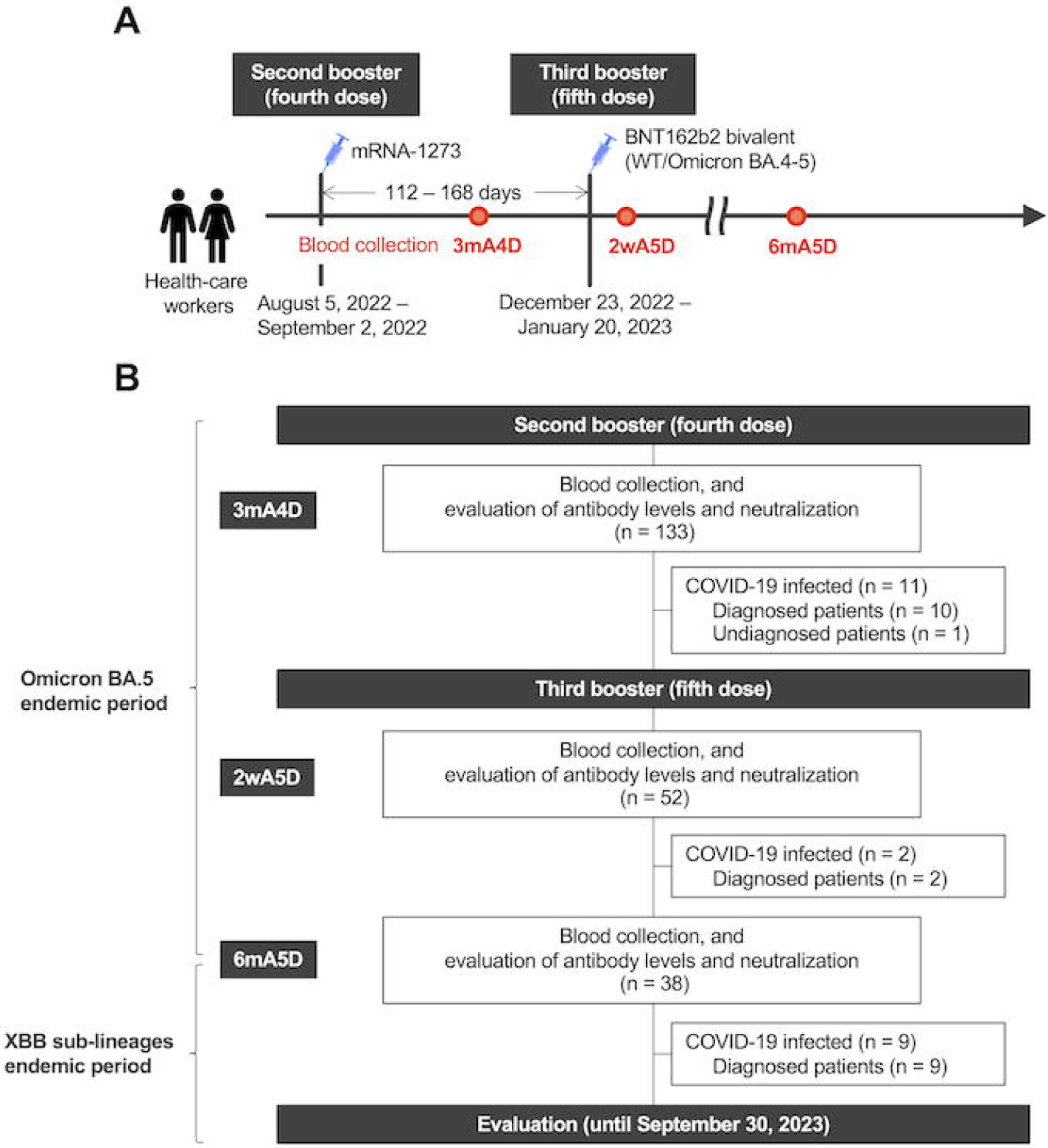
Overview of the study. **(A)** A schematic illustration of the time course of the study. **(B)** Study flow chart. Participants (n = 133) who received four monovalent vaccines provided blood samples, and their vaccine-induced antibody responses were assessed at 3mA4D. They subsequently received a fifth dose of the bivalent vaccine (WT/Omicron BA.4-5) and followed up at 2wA5D and 6mA5D. The occurrences of diagnosed and undiagnosed infections in participants were also followed and recorded. 3mA4D, 3 months after the fourth dose; 2sA5D, 2 weeks after the fifth dose; 6mA5D, 6 months after the fifth dose; WT, wild-type.

**Table 1.** Demographic characteristics of the study participants at the 3mA4D, 2wA5D, and 6mA5D.

| Profile | 3mA3D, n = 133 | 2wA5D, n = 52 | 6mA5D, n = 38 |
| --- | --- | --- | --- |
| Sex, male, n (%) | 35 (26.3) | 17 (32.7) | 12 (31.6) |
| Age, yrs, n (%) |  |  |  |
| 20–24 | 9 (6.8) | 3 (5.8) | 2 (5.3) |
| 25–29 | 11 (8.3) | 2 (3.8) | 1 (2.6) |
| 30–34 | 13 (9.8) | 3 (5.8) | 3 (7.9) |
| 35–39 | 16 (12.0) | 8 (15.4) | 7 (18.4) |
| 40–44 | 17 (12.3) | 6 (11.5) | 2 (5.3) |
| 45–49 | 21 (15.8) | 7 (13.5) | 6 (15.8) |
| 50–54 | 13 (9.8) | 5 (9.6) | 2 (5.3) |
| 55–59 | 19 (14.3) | 10 (19.2) | 8 (21.1) |
| 60–64 | 13 (9.8) | 7 (13.5) | 6 (15.8) |
| ≥65 | 1 (0.8) | 1 (1.9) | 1 (2.6) |

During the follow-up period, 10, 2 and 9 participants were diagnosed with COVID-19 between 3mA4D and 2wA5D, 2wA5D and 6mA5D (Omicron BA.5 predominant period), and after 6mA5D (XBB sub-lineages predominant period), respectively. The diagnosed infections were confirmed to be anti-N antibody positive. In one participant without self-reported history of COVID-19, the anti-N antibody was negative at 3mA4D, but became positive at 2wA5D; this was defined as an undiagnosed COVID-19 infection occurring between 3mA4D and 2wA5D (**Fig. 1**).

### Pre-infection humoral immune status in COVID-19 BIs

Among 11 participants who developed BIs between 3mA4D and 2wA5D (Omicron BA.5 predominant period), the ranges of time from the fourth dose of vaccination and the evaluation at 3mA4D to the symptom onset in the participants with diagnosed BI were 100–151 and 16–51 days, respectively. Pre-infection (at 3mA4D) anti-RBD antibody and NTs against the Omicron BA.5 variant were compared between the BI group (n = 11) and the control group (n = 122) of participants who did not develop a BI (**Fig. 2A and 2B**). The anti-RBD antibody levels in the BI group (median, 8324.0 U/L [interquartile range (IQR), 4995.0––14702.0 U/L]) were not significantly different from those in the control group (median, 12644.0 U/L [IQR, 7801.0–19393.0 U/L]) (**Fig. 2A**). The pre-infection htCRNT values against BA.5 obtained using 200-fold diluted sera in the BI group (median, 0.0% [IQR, 0.0%–18.8%]) were significantly lower than those in the control group (median, 18.9% [IQR, 0.0%–54.4%]) (**Fig. 2B**). Significant differences were also observed when using 100- and 400-fold diluted sera (**Fig. 2B**).

**Figure 2.**
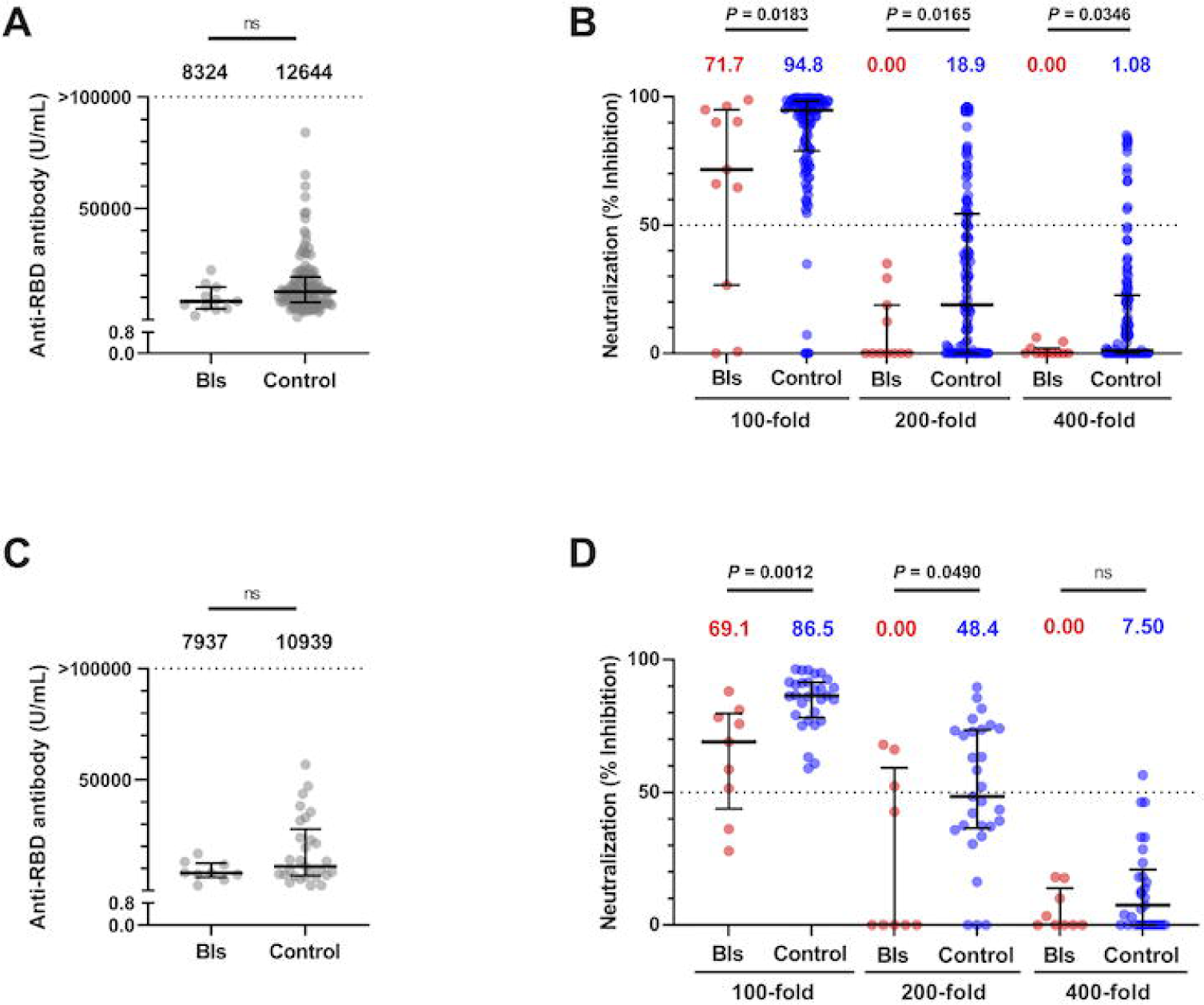
Pre-infection humoral immune levels in COVID-19 BI. **(A**) Serum concentration of anti-RBD antibody at 3mA4D (BA.5 endemic period) in the BI group (BIs, n = 11) and the controls (n = 122). Each dot represents an individual result. **(B)** Pseudotypevirus-based neutralizing activity against Omicron BA.5 at 3mA4D in the BI group (BIs, n = 11) and the controls (n = 122). **(C)** Serum concentrations of anti-RBD antibody at 6mA5D (XBB sub-lineages endemic period) in the BI group (BIs, n = 9) and the controls (n = 29). **(D)** Pseudotypevirus-based neutralizing activity against Omicron XBB.1.5 at 6mA5D in the BI group (BIs, n = 9) and the controls (n = 29). The assay was performed using 100-, 200-, and 400-fold diluted serum. The numbers at the top indicate the median neutralizing values of each group. RBD, receptor-binding domain; BIs, breakthrough infections; 3mA4D, 3 months after the fourth dose; 6mA5D, 6 months after the fifth dose; ns, not significant. Bars indicate medians with interquartile ranges.

Between 2wA5D and 6mA5D (Omicron BA.5 predominant period), we were unable to adequately compare between the BI and control groups due to the limited number of BIs. Only two participants developed BIs, and the time from the fifth dose of vaccination to the symptom onset was 19 and 23 days, respectively. The pre-infection (at 2wA5D) htCRNT values against BA.5 and XBB.1.5 obtained using 200-fold diluted sera were 0% and 58.6%, and 0% and 16.9%, respectively.

After evaluation at 6mA5D (Omicron XBB sub-lineages predominant period), 9 participants developed BIs. The ranges of time from the fifth dose of BA.4-5 bivalent vaccine and the evaluation at 6mA5D to the symptom onset in participants with diagnosed BI were 136–273 and 11–123 days, respectively. Pre-infection (at 6mA5D) anti-RBD antibodies and NTs against the Omicron XBB.1.5 variant were compared between the BI group (n = 9) and the control group (n = 29) of participants that did not develop a BI (**Fig. 2C and 2D**). The anti-RBD antibody levels in the BI group (median, 7937.0 U/L [IQR, 6060.0–12310.0 U/L]) were not significantly different from those in the control group (median, 10939.0 U/L [IQR, 6740.0–27738.0 U/L]) (**Fig. 2C**). The pre-infection htCRNT values against XBB.1.5 determined using 200-fold diluted sera in the BI group (median, 0.0% [IQR, 0.0%–59.2%]) were significantly lower than those in the control group (median, 48.4% [IQR, 36.5%–73.5%]) (**Fig. 2D**). Significant differences were also observed when using 100-fold diluted sera, but not when using 400-fold diluted sera due to the overall low htCRNT values (**Fig. 2D**).

Based on these findings and the assumption that htCRNT values were even lower at the time of infection than at the pre-infection evaluation, the htCRNT values below 50% determined using the 200-fold diluted sera (i.e., the pseudotyped virus-based half-maximal neutralizing titer [NT_50_] < 200) might have been one of the indicators of high risk for COVID-19 BI.

### Antibody quantification and neutralizing activity before and after the third booster

We previously reported the immunogenicity and safety of the BNT162b2 first booster (third dose) and mRNA-1273 second booster vaccines (fourth dose) at 2 weeks after the third dose (2wA3D), 3 months after the third dose (3mA3D), 6 months after the third dose (6mA3D), and 2 weeks after the fourth dose (2wA4D) in the same participants as in the present study [16]. In the present study, we assessed the durability of the antibodies and their NTs after the second and third boosters.

At 3mA4D, 2wA5D, and 6mA5D, the median concentration of anti-RBD antibody was 12216.0 U/mL (IQR, 7383.0–18447.0 U/mL), 22537.0 U/mL (IQR, 14703.0–44950.0 U/mL), and 9287.0 U/mL (IQR, 6882.0–21695.0 U/mL) (**Fig. 3A**). After the third booster dose of BA.4-5 bivalent vaccine, the anti-RBD antibodies at 2wA5D were significantly increased compared with those at 3mA4D; however, these antibodies decreased significantly over time, and at 6mA5D their levels were similar to those at 3mA4D.

**Figure 3.**
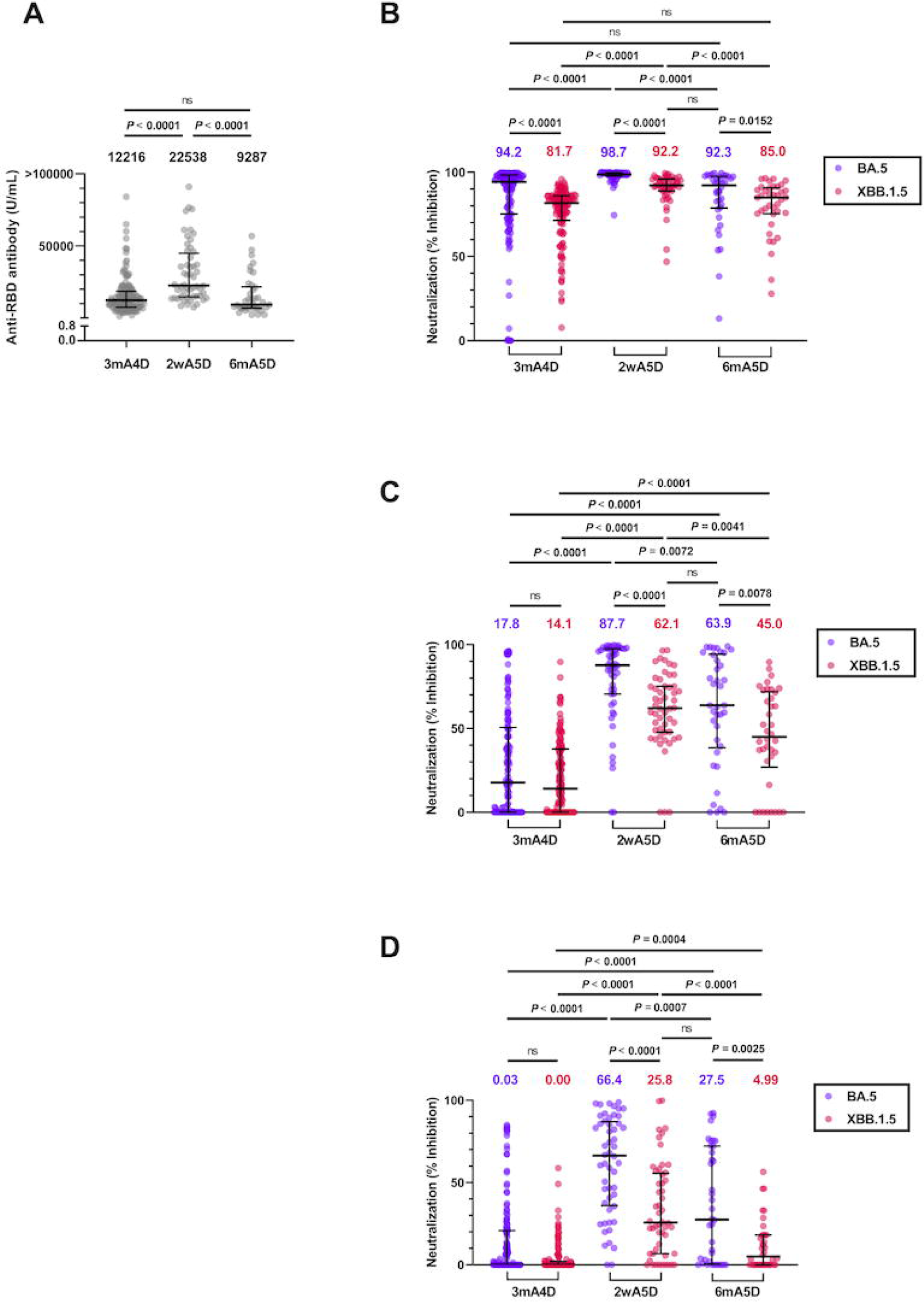
Anti-RBD antibody levels and neutralizing activities before and after the third booster. (A) Serum concentrations of anti-RBD antibody at 3mA4D (n = 133), 2wA5D (n = 52), and 6mA5D (n = 38). **(B–D)** Pseudotypevirus-based neutralizing activity against Omicron BA.5 and XBB.1.5 at 3mA4D (n = 133), 2wA5D (n = 52), and 6mA5D (n = 38). The assay was performed using **(B)** 100-, **(C)** 200-, and **(D)** 400-fold diluted serum. The numbers at the top indicate the median neutralizing values of each group. RBD, receptor-binding domain; 3mA4D, 3 months after the fourth dose; 6mA5D, 6 months after the fifth dose; ns, not significant. Bars indicate medians with interquartile ranges.

The median htCRNT values for Omicron BA.5 determined using the 100-, 200-, and 400-fold diluted sera at 3m4D were 94.2% (IQR, 75.2%–98.4%), 17.8% (IQR, 0.0%–50.7%), and 0.0% (IQR, 0.0%–20.8%) (**Fig. 3B, 3C, and 3D**). At 2wA5D, the median htCRNT values against BA.5 obtained using 100-, 200-, and 400-fold diluted sera (98.7% [IQR, 97.8%–99.8%], 87.7% [IQR, 70.6%–97.6%], and 66.4% [IQR, 36.1%–87.1%], respectively) were significantly increased compared with those at 3mA4D, but subsequently decreased at 6mA5D (92.3% [IQR, 78.8%–97.6%], 63.9% [IQR, 38.5%–94.2%], and 27.5% [IQR, 0.5%–72.2%], respectively) (**Fig. 3B, 3C, and 3D**).

Similar trends were observed in the htCRNT values against XBB.1.5; however, the htCRNT values against XBB.1.5 were significantly lower than those against BA.5 at 3m4D using 100-, 200-, and 400-fold diluted sera, and at 2wA5D and 6mA5D using 200- and 400-fold diluted sera (**Fig. 3B, 3C, and 3D**). The htCRNT values against XBB.1.5 at 2wA5D were similar to those against BA.5 at 6mA5D irrespective of whether 100-, 200-, or 400-fold diluted sera were used. In addition, although the anti-RBD antibodies at 3mA4D were not significantly different from those at 6mA5D (**Fig. 3A**), the htCRNT values against BA.5 and XBB.1.5 at 6mA5D were significantly higher than those at 3mA4D, when using either 200-fold or 400-fold diluted sera (**Fig. 3C and 3D**).

These results showed that, although the anti-RBD antibody levels were similar, NTs against not only BA.5 but also XBB.1.5 induced by the third booster dose of BA.4-5 bivalent vaccine were higher than those induced by the second booster dose of monovalent vaccine at 3mA4D, and were sustained even after 6 months.

### Correlation of neutralizing activity between DBS eluates and paired sera

To determine whether filter paper can be used to store human blood for serological testing, we tested DBS eluates in neutralization assays. DBS cards and paired sera were collected from individuals enrolled into this prospective observational study at the Toyama University Hospital. DBS cards were self-prepared via fingerstick or by spotting venous blood onto cards immediately after venipuncture, and paired sera were collected on the same day.

We first estimated the volume of sera on each 4-mm diameter DBS disc (area: 12.56 mm^2^); a single DBS disc was punched from each DBS card for addition to one well of a 96-well plate (**Fig. 4A**). The volume of blood obtained via fingerstick using a 0.8-mm Medisafe-finetouch needle (Terumo, Tokyo) was approximately 15 μL by repeated measurements. Thus 15 μL of blood was absorbed on each DBS card (nos. 545, 590, and 424 paper [Advantec, Tokyo]), the area was measured, and the volume of blood absorbed per mm^2^ was calculated as 0.39–0.94 μL/mm^2^ for both DBS cards. When DBS discs are eluted in 100 μL of DMEM containing 10% heat-inactivated FBS, and assuming that the serum constitutes approximately 50% of the whole blood, the eluates are estimated as approximate 20-fold diluted sera.

**Figure 4.**
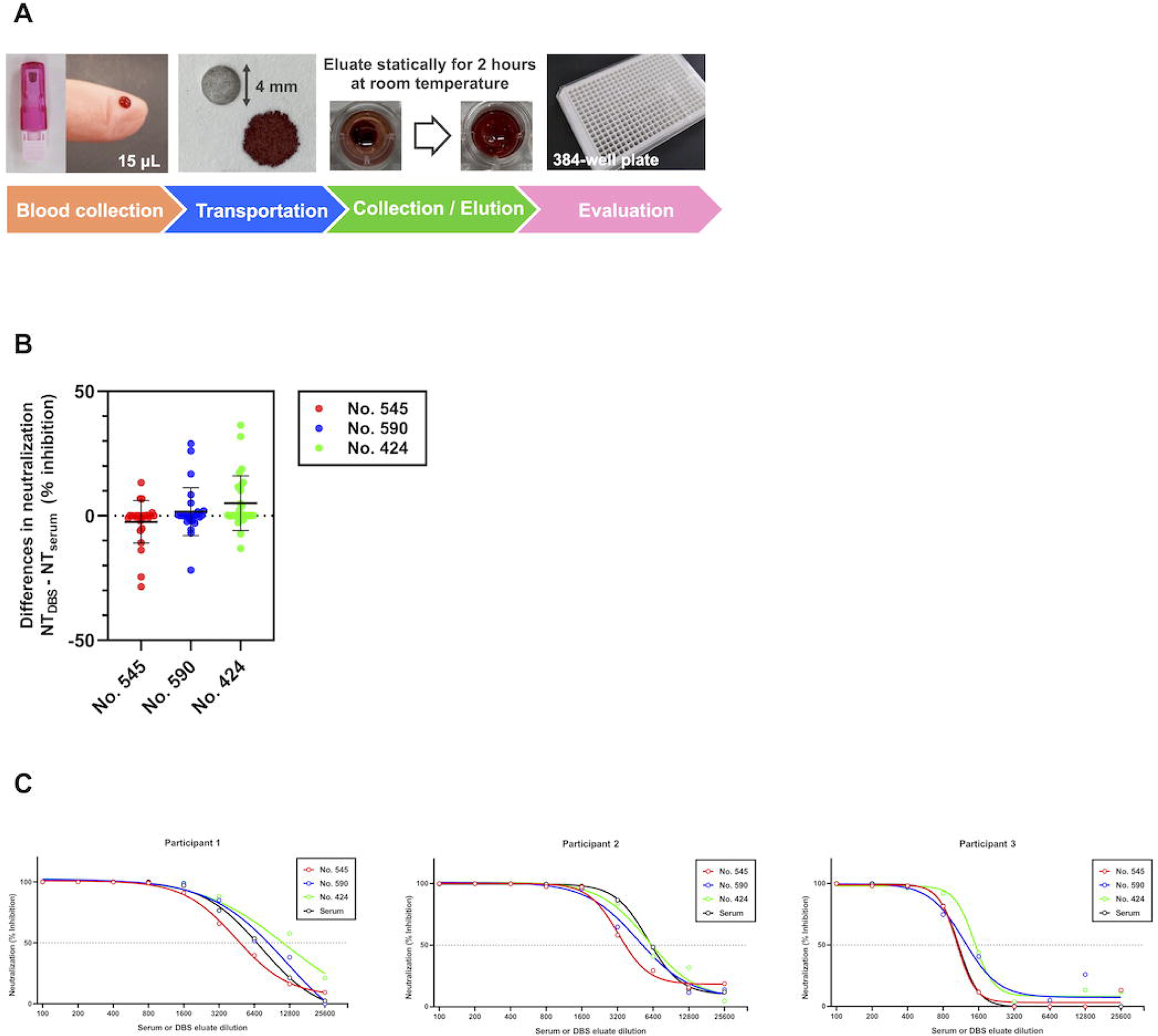
Evaluation of neutralizing activity using dried blood spot eluates. **(A**) Overview of the pseudotypevirus-based assay using dried blood spot (DBS) eluates. Approximately 15 μL of whole blood was obtained via fingerstick using a 0.8 mm Medisafe-finetouch needle (Terumo, Tokyo) and stored on each DBS card (no. 545, 590, and 424 filter papers [Advantec, Tokyo]) (i) at room temperature (RT) overnight (O/N); (ii) at RT for one month (1M); or (iii) at RT for 1M and then at 40°C for one additional month (2M/HT) before elution. For elution, a 4 mm disc was punched out from the center of each DBS card, and statically eluted with 100 μL of DMEM for 2 h at RT. DBS eluates were used for htCRNT [14, 16]. **(B)** The differences (NT_DBS_-NT_Serum_) in the htCRNT values against the WT evaluated using sera and DBS eluates of three vaccinated individuals. Three types of DBS cards (filter paper nos. 545, 590, and 424) were evaluated. The assay was performed using 100- to 25600-fold diluted serum and its equivalent, 5- to 1280-fold diluted DBS eluates. **(C)** The individual results for three participants. DBS, dried blood spot; RT, room temperature; O/N, overnight; 1M, one month at room temperature; 2M/HT, one month at room temperature followed by 40°C for one additional month; DMEM, Dulbecco’s modified Eagle’s medium; htCRNT, high throughput chemiluminescence reduction neutralization test; NT_DBS_, neutralizing activities using DBS eluates; NT_serum_, neutralizing activities using sera; WT, wild-type. Bars indicate mean with standard deviation.

To evaluate the correlation of NT between sera and DBS eluates, we evaluated the htCRNT values against the WT virus using sera (NT_serum_) and DBS eluates (NT_DBS_) at 100- to 25600-fold dilution in three vaccinated individuals. From the center of DBS cards, 4 mm discs were punched and statically eluted with 100 μL of DMEM for 2 h at room temperature (RT) one day after collections. As a result, regardless of the type of filter paper used, there was little difference in the htCRNT values against the WT virus between the DBS eluates and the paired sera at 100- to 25600-fold dilutions (**Fig. 4B**). The mean ± SD differences (NT_DBS_ - NT_Serum_) of the htCRNT values (%) were −2.5 ± 8.5% when using the no. 545, 1.7 ± 9.6% when using the no. 590, and 5.1 ± 11.0% when using the no. 424 filter paper (**Fig. 4B**). The individual results are shown in **Fig. 4C**.

To determine whether DBS eluates could be used to evaluate the htCRNT values against other variants and to evaluate the time and temperature stability, we then compared the htCRNT values against BA.5 or XBB.1.5 between the DBS eluates and paired sera (at 200-fold dilution) collected at 6mA5D in 12 vaccinated individuals, respectively. For evaluation of time and temperature stability, DBS cards were stored at RT for one month (1M) and thereafter at 40°C for one more month (2M/HT) before elution. The htCRNT values against BA.5 using both 10-fold diluted 1M- and 2M/HT-DBS eluates (equivalent to 200-fold diluted sera) were significantly correlated with the anti-RBD antibody levels and those using 200-fold diluted sera (**Fig. 5A, 5B, 5C, and 5D**). On the other hand, the htCRNT values against XBB.1.5 using 10-fold diluted 1M-DBS eluate were significantly correlated with the levels of anti-RBD antibodies and those using 200-fold diluted paired sera on filter papers no. 545 and 424, but not 590 (**Fig. 5E and 5F**). When using 2M/HT DBS eluates, the htCRNT values against XBB.1.5 on filter paper no. 545 were significantly correlated with the anti-RBD antibody levels and those using 200-fold diluted paired sera, but these correlations were not observed for filter papers no. 590 and 424 (**Fig. 5G and 5H**).

**Figure 5.**
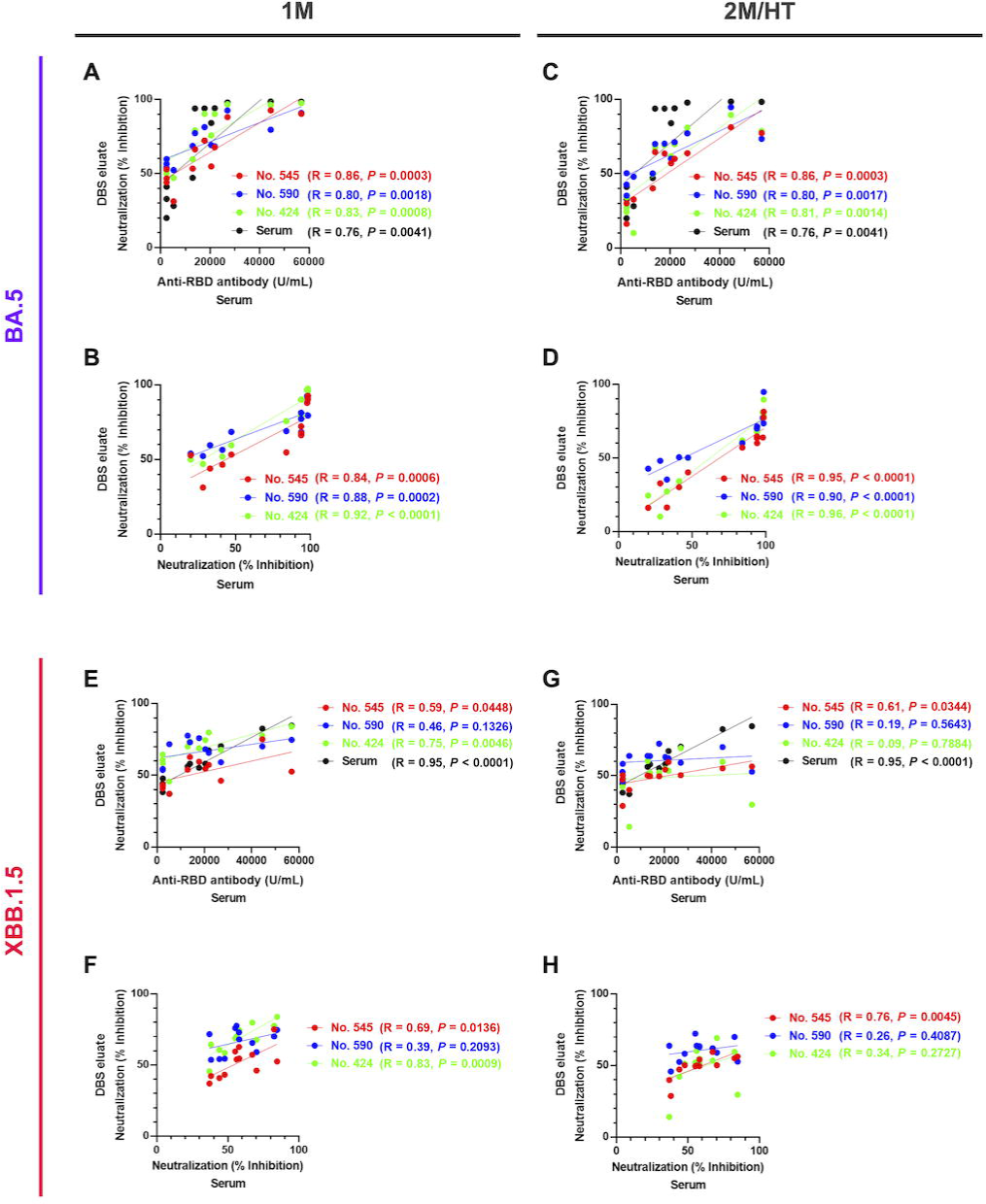
Correlation of neutralizing activities against BA.5 and XBB.1.5 between DBS eluates and paired sera. Relationship between htCRNT levels against BA.5 determined using 10-fold dilution of DBS eluates at **(A)** 1M or **(C)** 2M/HT and anti-RBD antibody levels. Relationship between htCRNT levels against BA.5 determined using 10-fold dilution of DBS eluates **(B)** at 1M or **(D)** at 2M/HT and its equivalent, 200-fold dilution of paired sera. Relationship between htCRNT levels against XBB.1.5 determined using 10-fold dilution of DBS eluates at **(E)** 1M or **(G)** 2M/HT and anti-RBD antibody levels. Relationship between htCRNT levels against XBB.1.5 determined using 10-fold dilution of DBS eluates **(F)** at 1M or **(H)** at 2M/HT and its equivalent, 200-fold dilution of paired sera. DBS, dried blood spot; RBD, receptor-binding domain; 1M, one month at room temperature; 2M/HT, one month at room temperature followed by 40°C for one additional month.

## DISCUSSION

We found that participants who went on to develop a BI had lower levels of NTs in the pre-infection period compared to participants that did not develop a BI. This difference was consistently observed both in the BA.5 and XBB sub-lineage endemic periods, but only with respect to NTs; the difference in anti-RBD antibodies was not statistically significant. The present study also showed that the htCRNT values below 50% using the 200-fold diluted sera (NT_50_ < 200) might be one of the indicators of high risk for COVID-19 BIs. These results were consistent with the previous studies [9, 23]. In the study conducted to assess the humoral response in a population of healthcare workers who received three doses of the BNT162b2 mRNA COVID-19 vaccine [9], pseudotyped virus-based half-maximal neutralizing titers (NT_50_) against Omicron BA.1, but not total binding antibodies, were significantly lower and all below 200 in subjects that developed BI with Omicron BA.1. Another study conducted around the time of the BA.5 endemic wave, approximately 6 months after participants received a third dose of COVID-19 mRNA vaccine, also showed that titers (NT_50_) evaluated using live virus assay were significantly lower in the BI cases than in controls [23]. To the best of our knowledge, this is the first study to demonstrate that these differences were also observed after a fourth dose of monovalent and fifth dose of bivalent (WT/BA.4-5) vaccine, and during not only the BA.5 but also the XBB sub-lineage endemic periods. In addition, a protection threshold of NT_50_ < 200 would be important for an individual’s protection against infection, and this threshold could also be valuable in considering social vaccination strategies.

The present study also showed that the bivalent (WT/BA.4-5) vaccine elicited higher levels of neutralizing antibodies against Omicron XBB.1.5, but similar to previous studies [24, 25], the htCRNT values against XBB.1.5 were significantly lower than those against BA.5. Actually, the htCRNT values against XBB.1.5 at 2wA5D were similar to those against BA.5 at 6mA5D. Previous studies using live virus assay also demonstrated that the gMean-NT_50_ values against XBB1.5 at 2wA5D of bivalent (WT/BA.4-5) vaccine were 61 (range: <20-−2175) [24], equivalent to those against BA.5 at 24 weeks after the fifth dose of bivalent (WT/BA.4-5) vaccine (74 [<20–1089]) [25]. The present study demonstrated that the median htCRNT value using 200-fold diluted sera against XBB.1.5 at 6mA5D was 45%, indicating that almost half of the infection-naïve participants are at high risk of BI with XBB.1.5. This underscores the importance of the Omicron XBB.1.5 monovalent vaccine booster.

An easily implementable serological assay to accurately assess neutralizing antibodies is also urgently needed to better track herd immunity, vaccine efficacy, and individual immune status, especially during pandemics with restrictions on direct patient contact. Although SARS-CoV-2 antibody testing using serum or plasma is reliable and widely used, the sample collection poses several logistical restrictions, such as the requirements of venipuncture by trained phlebotomists with direct contact, immediate storage in a refrigerator or freezer after collection, and cold chain transport to maintain the biospecimen integrity. DBS overcomes these barriers, as the samples can be self-collected by fingerstick, and then mailed and stored at ambient temperature [19]. We demonstrated that neutralizing antibody responses measured using DBS eluates correlated with those measured from paired sera. A related study conducted by Itell et al. also provided support for the use of DBS to measure neutralizing antibody responses by pseudotyped virus assay [19]; however, only the neutralizing antibody against the WT strain was assessed [19].

DBS sampling also provides a means of widening access to humoral immunity evaluation in remote areas of low- and middle-income countries. Although some studies have shown that DBS eluates retained comparable neutralizing antibody responses to paired sera after storage on filter paper for 28 days at RT [13], RT can vary in different settings, with developing countries often experiencing higher RT conditions. To the best of our knowledge, the effect of higher temperatures on neutralizing antibodies has not been elucidated.

The present study is the first to demonstrate the correlations between neutralizing antibody responses against not only the WT strain but also BA.5 and XBB.1.5 in the DBS eluate and paired sera. In addition, neutralizing antibodies have been shown to remain stable in collected DBS transported at HT(40°C) for one month following one-month storage at RT. However, in our experiments the correlations and time and temperature stability differed according to the type of filtered papers used, with no. 545 filter paper being the most suitable for DBS cards in the htCRNT assay. Neutralizing antibody evaluation using DBS eluates could be applied to other pathogens and in future pandemics, facilitating reliable and affordable seroepidemiological surveillance, including in remote areas and low-income countries [26].

There are several limitations of this study that need to be addressed in future research. First, we included only participants aged 20 to 69 years, and simplified background information was collected [16]. Thus, the antibody responses in older and younger individuals are unknown. In addition, since detailed interviews regarding specific underlying diseases and medications were not conducted, it was impossible to infer the impact of the health of individual participants [16]. Second, the participants infected after 6mA5D were not confirmed to be anti-N antibody-positive, and we did not perform genomic analysis of the infected variants in participants with BIs. Third, we did not evaluate neutralizing antibody responses using a live virus assay, although in our previous study we demonstrated the strong correlations between the htCRNT and live virus assays [16]. We considered that a complex live virus assay using DBS eluates would be beyond the scope of this study, which was focused on establishing an easily implementable serological assay using DBS cards. Finally, although we evaluated the application of DBS sample collection with three commonly used filter papers (nos. 545, 590, and 424), we cannot rule out that other more suitable filter papers may exist.

In conclusion, our results showed that the participants who developed BI had lower levels of neutralizing antibody, but no difference in anti-RBD antibody levels compared to the controls without BI. These differences were consistently observed after the fourth dose of monovalent and fifth dose of bivalent (WT/BA.4-5) vaccine, and during not only BA.5 but also XBB sub-lineage endemic periods. Given the above results, the htCRNT values below 50% using the 200-fold diluted sera might be one of the indicators of high risk of COVID-19 BI. In almost half of infection-naïve participants, the NTs against XBB.1.5 at 6mA5D were lower than this threshold, suggesting the need for XBB.1.5 monovalent vaccination. Furthermore, DBS sample collection could make it easier to determine the immune status of individual patients against the rapidly changing endemic variants, since it has the advantages of less invasive collection (i.e., self-collection), low cost, and easy transport and storage, making it a practical alternative to blood collection via traditional venipuncture. These advantages would also make DBS appealing for use in resource-limited settings and in potential future pandemics.

## Transparency declaration

### Data availability

All data are provided in the manuscript and supplementary information.

## Acknowledgments

We thank Masako Aoki, Misato Matsuura, and Shiori Sasahara for their contribution to data collection and the staff at Toyama University Hospital for their help in collecting the specimens and questionnaires.

## Author contributions

Conceptualization: H.K. and Y. Morinaga; Methodology: H.K., Y. Morinaga, and H.T.; Validation: H.K. and Y. Morinaga; Formal Analysis: H.K. and Y. Morinaga; Investigation: H.K., Y. Morinaga, H.Y., Y. Yoshida, M.E., Y.K., Y.T., M.K., Y. Murai, K.K. (neutralizing assay), and H.N. (commercial antibody test); Resources: H.T. (generating pseudotyped viruses), H.K., M.E., Y.K., Y.T., M.K., Y. Murai, K.K., and K.N. (serum sample collection); Data Curation: H.K. and Y. Morinaga; Writing–Original Draft Preparation: H.K., Y. Morinaga and H.T.; Writing–Review and Editing: H.K., Y. Morinaga, and H.T.; Visualization: H.K. and Y. Morinaga; Supervision: Y. Morinaga, H.N., and Y. Yamamoto; Project Administration: Y. Morinaga and Y.Yamamoto; Funding acquisition: H.K., Y. Morinaga, H.T., H.N., and Y. Yamamoto.

## List of abbreviations

SARS-CoV-2: Severe acute respiratory syndrome coronavirus 2
COVID-19: Coronavirus disease 2019
mRNA: Messenger RNA
VE: Vaccine efficacy
WT: Wild-type
BI: Breakthrough infection
htCRNT: High throughput chemiluminescence reduction neutralization test
DBS: Dried blood spots
RBD: Receptor-binding domain
NT: Neutralizing activity
3mA4D: 3 months after the fourth dose
2wA5D: 2 weeks after the fifth dose
6mA5D: 6 months after the fifth dose
N: SARS-CoV-2 nucleocapsid
RT: room temperature
HT: high temperature
1M: one month at room temperature
2M/HT: one month at room temperature followed by 40°C for one additional month
DMEM: Dulbecco’s modified Eagle’s medium
FBS: fetal bovine serum
VSVs: Vesicular stomatitis viruses
SD: Standard deviation
IQR: Interquartile range
NT_50_: Half-maximal neutralizing titer
2wA3D: 2 weeks after the third dose
3mA3D: 3 months after the third dose
6mA3D: 6 months after the third dose
2wA4D: 2 weeks after the fourth dose
NT_serum_: neutralizing activities using sera
NT_DBS_: neutralizing activities using dried blood spot eluates

